# Metabolic Phenotypes of Overweight/Obesity and Risk of Cardiovascular Diseases in Postmenopausal Women: Findings from the China PEACE Million Persons Project

**DOI:** 10.1101/2023.09.25.23296122

**Authors:** Danying Deng, Zhiqiang Nie, Jiabin Wang, Chaolei Chen, Wenbin Wang, Yanchen Zhu, Qingyu Guan, Yanqiu Ou, Yingqing Feng

**Author notes:** **Correspondence to**: Yingqing Feng, PhD, Guangdong Provincial Key Laboratory of Coronary Heart Disease Prevention, Guangdong Cardiovascular Institute, Guangdong Provincial People’s Hospital (Guangdong Academy of Medical Sciences), Southern Medical University, Guangzhou 510080, China.

## Abstract

**BACKGROUND:** Obesity and metabolic abnormalities were associated with increased risk of cardiovascular diseases (CVD). However, much remains unknown about which metabolic weight phenotypes and how relate to CVD, especially in postmenopausal women.

**METHODS AND RESULTS:** We included 15,575 postmenopausal women aged 35 to 75 years (median age, 60.6 [IQR: 55.0-65.7]) free of CVD at baseline from a subcohort of the China Patient-centered Evaluative Assessment of Cardiac Events Million Persons Project between 1 January 2016 and 31 December 2020 in Guangdong Province at 8 sites. Cox regression models were used to investigate the associations of metabolic and overweight/obese status with the risk of developing CVD. Over a median follow-up period of 3.32 years, a total of 1354 CVD events occurred. Overall, 30.3% were metabolically heathy among the 9404 overweight/obese participants, while 49.2% were metabolically unhealthy among the 6171 normal weight subjects. Compared with the metabolically healthy normal weight group, the remaining three groups of participants were all at higher risk of combined CVD, with multivariate-adjusted hazard ratios and 95% confidence intervals of 1.41 (1.16–1.71) for metabolically unhealthy normal weight, 1.42 (1.16–1.73) for metabolically healthy overweight/obesity, and 1.75 (1.47–2.07) for metabolically unhealthy overweight/obesity. When subdividing overweight/obesity separately into overweight and obesity, combined CVD risk was only greater in metabolically unhealthy individuals across different weight categories.

**CONCLUSIONS:** Postmenopausal women with metabolically healthy overweight/obesity and metabolically unhealthy normal weight had a higher risk of cardiovascular disease than those with metabolically healthy normal weight, and the association of metabolically unhealthy overweight/obesity was greatest among four categories, suggesting that metabolic abnormalities and overweight/obesity should be monitored and controlled.

**CLINICAL PERSPECTIVE:** **What Is New?**

- Postmenopausal women with metabolic abnormalities combined with overweight/obesity and even normal weight had a higher risk of cardiovascular disease (CVD) than those with metabolically healthy normal weight.
- Metabolically healthy overweight/obesity were associated with greater risk of CVD, indicating that overweight/obesity alone may contribute to CVD risk in postmenopausal women.

**What Are the Clinical Implications?**

- Individuals who are metabolically unhealthy and overweight/obese warrant close clinical supervision, while those of normal weight should not be ignored.

---

Cardiovascular diseases (CVDs) are the leading cause of mortality and generate an enormous health burden worldwide,^1^ and they were responsible for 35% of total deaths in women in 2019.^2^ Overall, the prevalence of CVD grows rapidly in women when reaching 50 years or older and tends to exceed that of men.^3^ Obesity increases cardiovascular risk factors, leading to metabolic abnormalities (also known as metabolic syndrome) as well as endocrine disorders (including dyslipidemia and insulin resistance) and therefore promoting hypertension and diabetes.^4^

The risk of CVD was shown to increase as body mass index (BMI) increases,^5^ and metabolic abnormalities (e.g., elevated blood pressure, hyperglycemia) were reported to be related to CVD.^6^ Nevertheless, not all overweight/obese individuals suffer from metabolic abnormalities, and some people with metabolic syndrome are not overweight/obese, separately known as metabolically healthy overweight/obesity (MHO) and metabolically unhealthy normal weight (MUHNW). Previous studies showed that both MHO and MUHNW phenotypes were associated with a higher CVD risk;^7–10^ however, some research indicated only MHO^11^, and others suggested that only MUHNW^12–14^ was associated with CVD risk when compared with metabolically healthy normal weight individuals. These inconsistent results demonstrate that the correlations between CVD with MHO and MUHNW phenotypes remain controversial. Given the differences in CVD prevalence between the sexes and the fact that few studies have focused on females, we principally investigated women after menopause. Therefore, the purpose of this study was to examine the relationship between metabolically healthy/unhealthy status of overweight/obesity and incident CVD in postmenopausal women in China.

## METHODS

### Data Availability Statement

The original contributions are presented in the article/supplementary material, further inquiries can be directed to the corresponding author.

### Study Participants

We analysed data from a subcohort of the China PEACE MPP (Patient-centered Evaluative Assessment of Cardiac Events Million Persons Project) in Guangdong Province, which was a large-scale screening program funded by the government for identifying individuals at high risk of CVD.^15^ The China-PEACE MPP design and methods have been described previously.^15–17^ In total, 102 358 participants aged 35 to 75 years from 8 sites with local residence registration were enrolled between 1 January 2016 and 31 December 2020. Among them, a total of 16 381 women were postmenopausal. In the current study, we excluded participants with CVD at baseline (n=325) and those who were underweight, defined as (body mass index) BMI <18.5 kg/m^2^ (n=481). Finally, 15 575 postmenopausal women were included for analysis (**Figure S1**). Each participant provided signed informed consent. The present research was approved by the Ethics Review Committee of Guangdong Provincial People’s Hospital.

### Assessment of Covariates

Physical examinations were conducted to measure height, weight, systolic blood pressure (SBP), diastolic blood pressure (DBP), and waist circumference (WC). BMI was calculated by dividing the weight in kilograms by the square of height in meters. Blood pressure (BP) was measured twice on the right upper arm using an electronic monitor (HEM-7430, Omron Corporation, Kyoto, Japan). Fingertip blood samples were collected to determine total cholesterol (TC), triglyceride (TG), high-density lipoprotein cholesterol (HDL-C), and fasting blood glucose (FBG) from all subjects on an empty stomach. Standardized questionnaires were designed to gather sociodemographic information (age, sex, education level, marital status, household annual income, residence, and occupation), lifestyle behaviors (current smoking and alcohol consumption status), medications (use of antihypertensive, hypoglycemic, antiplatelet agents, and statins), and personal history of disease (self-reported hypertension, diabetes, and dyslipidemia). Diabetes was defined as FBG ≥ 7 mmol/L, self-reported physician diagnosis, or current use of hypoglycemic agents. Hypertension was defined as SBP/DBP ≥ 140/90 mmHg, self-reported history, or taking antihypertensive drugs. Dyslipidemia was defined as LDL-C ≥ 4.1 mmol/L, TC ≥ 6.2 mmol/L, self-reported history, or receiving lipid-lowering medications.

### Definitions of Obesity and Metabolic Phenotypes

Overweight/obesity was defined as BMI ≥ 24 kg/m^2^ or WC ≥ 85 cm, and normal weight was defined as BMI of 18.5-23.9 kg/cm^2^ and WC < 85 cm according to the Working Group on Obesity in China.^18, 19^ A metabolically unhealthy phenotype was defined as 2 or more of the following 4 abnormal components: (1) BP ≥ 130/85 mm Hg or current use of antihypertensive drugs, (2) FBG ≥ 5.6 mmol/L or current use of antidiabetic agents, (3) TGs ≥ 1.7 mmol/L, and (4) HDL-C < 1.3 mmol/L, according to the harmonized International Diabetes Federation criteria.^20^ Therefore, based on weight and metabolic status, the participants were classified into four groups: metabolically healthy normal weight (MHNW), metabolically unhealthy normal weight (MUHNW), metabolically healthy overweight/obesity (MHO), and metabolically unhealthy overweight/obesity (MUHO).

### Outcomes

Our primary outcome was incident CVD, a composite of incident events including stroke, heart failure (HF), coronary heart disease (CHD) and CVD death. The secondary outcomes of interest were separate stroke, HF and CHD. The codes of the Tenth Revision of International Classification of Diseases were used to identify the outcomes: I61–I64 for stroke, I50 for HF, and I20–I25 for CHD.^21^ The follow-up time was calculated by baseline, defined as the time of recruitment to the last follow-up date, 31 December 2021. All outcomes were collected from China’s Centre for Disease Prevention and Control’s National Mortality Surveillance System and Vital Registration.

### Statistical Analysis

Continuous variables are described as medians and interquartile ranges (IQRs) for nonnormally distributed data, and categorical variables are described as numbers (percentages). Differences in baseline characteristics among the four groups (MHNW, MUHNW, MHO, MUHO) were compared using the Kruskal–Wallis *H*-test or chi-square test as appropriate. The Kaplan–Meier method was used to estimate the cumulative incidence of cardiovascular outcomes in participants with different weights and metabolic statuses, and the log-rank test was used to compare differences in estimates. Cox proportional hazards models were applied to assess the associations between metabolic weight categories and the risk of cardiovascular events with the MHNW group as the reference, presenting hazard ratios (HRs) and 95% confidence intervals (CIs) as the results. Model 1 was adjusted for none. Model 2 was adjusted for age. Model 3 was fully adjusted for age, education level, marital status, family annual income, urban residence, farmer, current smoking, current alcohol consumption, total cholesterol, statin use, and antiplatelet medication.

According to participants who were metabolically healthy versus unhealthy, a sensitivity analysis was performed stratifying those classified as overweight/obesity separately into overweight and obesity in addition to normal weight, only based on BMI (6 groups total). We also examined the independent associations of each of the individual obesity and metabolic variables classified by cut points with incident CVD outcomes using adjusted Cox regressions, as well as the relationships between the number of abnormal metabolic components and CVD risk. Finally, we conducted stratification analyses by age (<65 vs. ≥65 years), education level (less than high school vs. high school or above), family annual income (≤50 000 vs. >50 000 China Yuan), and urban residence (no vs. yes), reporting *P* values for interactions.

The analyses above were conducted using IBM SPSS statistical software, version 26 (IBM Corporation, Armonk, New York, USA), and GraphPad Prism 8 (GraphPad Prism Software Inc., San Diego, CA, USA) was used to plot survival curves. A two-tailed *P* < 0.05 was considered statistically significant.

## RESULTS

### Baseline Characteristics

As illustrated in the study flow chart (**Figure S1**), a total of 15 575 postmenopausal women were included. The baseline characteristics of the participants compared among MHNW (20.1%), MUHNW (19.5%), MHO (18.3%), and MUHO (42.1%) are presented in **Table 1**, with a median age of 60.6 years. Of all participants, 60.4% were overweight/obese, and 61.6% were metabolically unhealthy. Among the 9404 overweight/obese subjects, 30.3% were metabolically healthy, while 49.2% were metabolically unhealthy among the 6171 normal weight subjects. Compared with metabolically healthy participants, metabolically unhealthy participants, whether obese or not, were older, less educated, more likely to be urban residents, and exhibited a much worse risk profile, including lower HDL-C and higher BMI, WC, SBP, DBP, TG, and FBG levels. BMI, WC and HDL-C were higher in the MHO than MUHNW. In addition, the MUHO group showed the highest levels of BMI, WC, SBP, DBP, TG and FBG compared with the other three groups.

**Table 1.** Baseline Characteristics According to Metabolic Weight Categories in Postmenopausal Women.

| Characteristic | MHNW | MUHNW | MHO | MUHO | *P* value |
| --- | --- | --- | --- | --- | --- |
| No. of participants | 3133 | 3038 | 2852 | 6552 |  |
| Age, years | 58.8 (53.7–64.0) | 61.0 (55.4–66.2) | 59.4 (54.3–64.9) | 61.5 (55.9–66.4) | <0.001 |
| Education level, No. (%) |  |  |  |  | <0.001 |
| Less than high school | 2438 (77.8) | 2417 (79.6) | 2433 (85.3) | 5589 (85.3) |  |
| High school or above | 695 (22.2) | 621 (20.4) | 419 (14.7) | 963 (14.7) |  |
| Married, No. (%) | 2712 (86.6) | 2626 (86.4) | 2487 (87.2) | 5710 (87.1) | 0.69 |
| Family annual income, No. (%) |  |  |  |  | 0.65 |
| ≤50 000 CNY | 1820 (58.1) | 1743 (57.4) | 1614 (56.6) | 3787 (57.8) |  |
| >50 000 CNY | 1313 (41.9) | 1295 (42.6) | 1238 (43.4) | 2765 (42.2) |  |
| Urban, No. (%) | 1358 (43.3) | 1464 (48.2) | 1079 (37.8) | 2810 (42.9) | <0.001 |
| Farmer, No. (%) | 423 (13.5) | 385 (12.7) | 344 (12.1) | 821 (12.5) | 0.39 |
| Current smoking, No. (%) | 21 (0.7) | 38 (1.3) | 23 (0.8) | 49 (0.7) | 0.047 |
| Current drinking, No. (%) | 28 (0.9) | 25 (0.8) | 30 (1.1) | 47 (0.7) | 0.42 |
| BMI, kg/m<sup>2</sup> | 21.6 (20.4–22.7) | 22.1 (20.9–23.0) | 25.5 (24.4–27.2) | 26.1 (24.6–28.1) | <0.001 |
| WC, cm | 76.0 (72.0–80.0) | 78.0 (74.0–81.0) | 87.0 (83.0–91.0) | 89.0 (85.0–94.0) | <0.001 |
| SBP, mmHg | 121.5 (111.5–131.0) | 139.5 (129.5–156.0) | 124.5 (116.5–137.0) | 142.5 (131.0–160.0) | <0.001 |
| DBP, mmHg | 72.5 (67.0–79.5) | 81.0 (73.5–88.0) | 75.0 (69.5–82.0) | 82.5 (75.0–90.0) | <0.001 |
| TC, mmol/L | 5.4 (4.6–6.3) | 5.4 (4.5–6.5) | 5.4 (4.6–6.3) | 5.4 (4.5–6.4) | 0.49 |
| TG, mmol/L | 1.1 (0.9–1.4) | 1.7 (1.2–2.3) | 1.2 (1.0–1.5) | 1.9 (1.4–2.6) | <0.001 |
| HDL-C, mmol/L | 1.8 (1.5–2.1) | 1.4 (1.2–1.8) | 1.7 (1.4–2.0) | 1.4 (1.1–1.6) | <0.001 |
| FBG, mmol/L | 5.2 (4.8–5.6) | 6.0 (5.6–6.7) | 5.2 (4.8–5.5) | 6.2 (5.6–7.1) | <0.001 |
| Hypertension, No. (%) | 738 (23.6) | 1848 (60.8) | 985 (34.5) | 4600 (70.2) | <0.001 |
| Diabetes, No. (%) | 181 (5.8) | 705 (23.2) | 232 (8.1) | 2003 (30.6) | <0.001 |
| Dyslipidemia, No. (%) | 969 (30.9) | 1125 (37.0) | 963 (33.8) | 2418 (36.9) | <0.001 |
| Anti-hypertensive medication, No. (%) | 45 (1.4) | 195 (6.4) | 47 (1.6) | 427 (6.5) | <0.001 |
| Hypoglycemic medication, No. (%) | 14 (0.4) | 83 (2.7) | 18 (0.6) | 158 (2.4) | <0.001 |
| Statin use, No. (%) | 18 (0.6) | 29 (1.0) | 18 (0.6) | 66 (1.0) | 0.08 |
| Anti-platelet medication, No. (%) | 4 (0.1) | 20 (0.7) | 5 (0.2) | 56 (0.9) | <0.001 |

Data are median (interquartile range) or number (%). MHNW indicates metabolically healthy normal weight; MUHNW, metabolically unhealthy normal weight; MHO, metabolically healthy overweight/obesity; MUHO, metabolically unhealthy overweight/obesity; CNY, China Yuan; BMI, body mass index; WC, waist circumference; SBP, systolic blood pressure; DBP, diastolic blood pressure; TC, total cholesterol; TG, triglyceride; HDL-C, high-density lipoprotein cholesterol; and FBG, fasting blood glucose.

### Metabolic Weight Categories and Risk of Cardiovascular Events

Over a median follow-up of 3.32 years, a total of 1354 (8.7%) composite cardiovascular events occurred, including 842 (5.4%) strokes, 518 (3.3%) CHD cases, 273 (1.8%) HF cases and 43 (0.3%) fatal cardiovascular events. The results of CVD death alone were not listed due to the limited number of events, and the associations were insignificant. Moreover, the risks of combined CVD among metabolic weight categories were consistent regardless of whether CVD death was considered. The Kaplan–Meier curves showed that higher cumulative incidences of combined CVD were observed among participants with MUHO, MHO, and MUHNW than those with MHNW (**Figure 1**).

**Figure 1.**
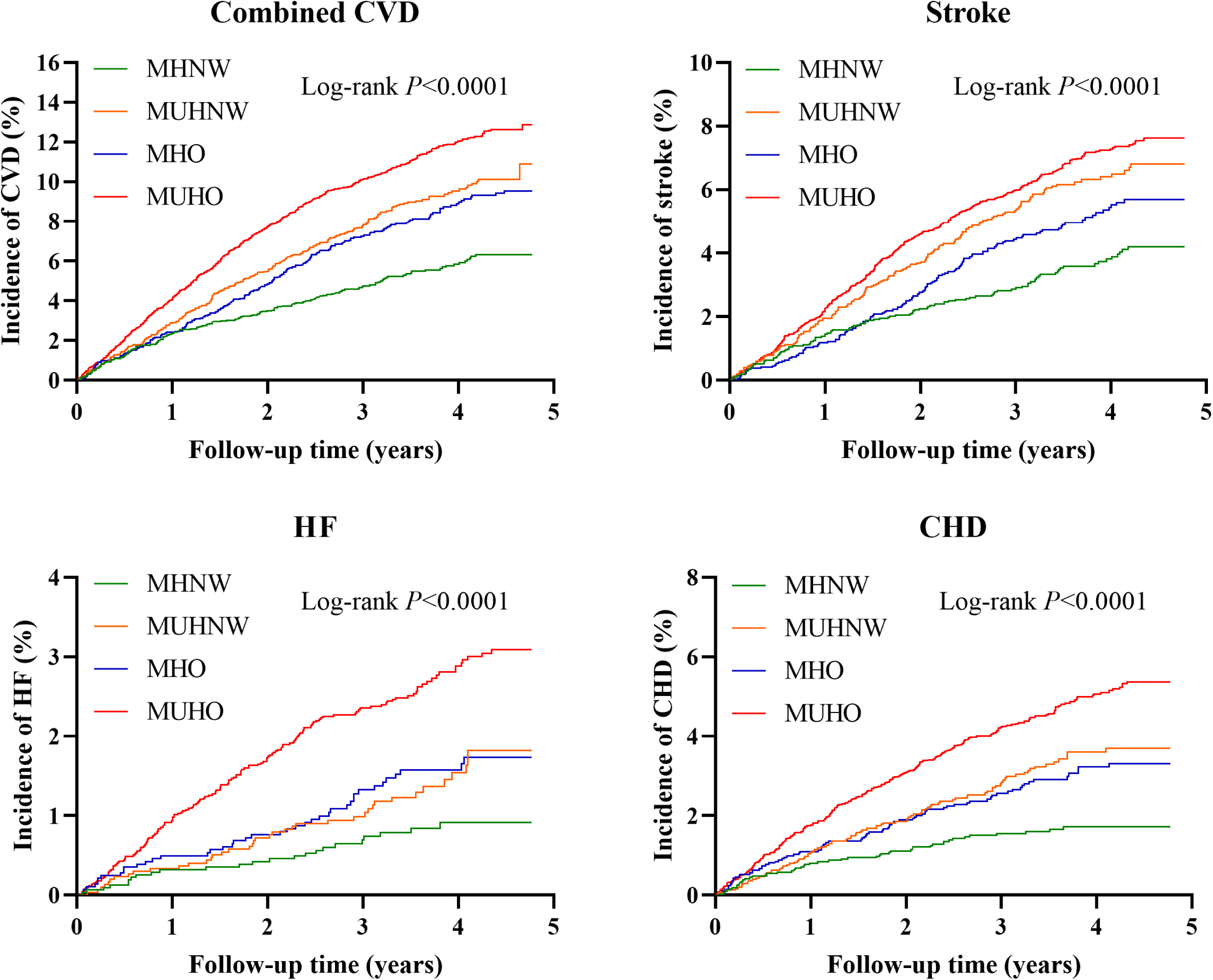
Unadjusted Kaplan–Meier curves for incident cardiovascular events stratified by metabolic weight categories. CVD indicates cardiovascular; HF, heart failure; and CHD, coronary heart disease.

As shown in **Table 2**, MUHNW was associated with a 41% increased risk of combined CVD (HR: 1.41; 95% CI: 1.16–1.71), MHO a 42% increased risk (HR: 1.42; 95% CI: 1.16–1.73), and MUHO a 75% increased risk (HR: 1.75; 95% CI: 1.47–2.07) compared with the MHNW group in the fully adjusted model. The multivariate-adjusted HRs with 95% CIs for stroke in the MUHNW, MHO and MUHO groups were 1.49 (1.17–1.89), [1.29 (1.00–1.67), *p* =0.047] and 1.60 (1.29–1.98), respectively. MHO (multivariate-adjusted HR = 1.75; 95% CI: 1.05–2.89) and MUHO [2.59 (1.69–3.98)] were associated with an increased risk of HF, whereas the association of MUHNW [1.38 (0.83–2.29)] was not significant. In the multivariate Cox regression model for CHD, the HRs with 95% CIs were 1.63 (1.15–2.30) for MHNW, 1.71 (1.20–2.44) for MHO, and 2.36 (1.74–3.20) for MUHO.

**Table 2.**
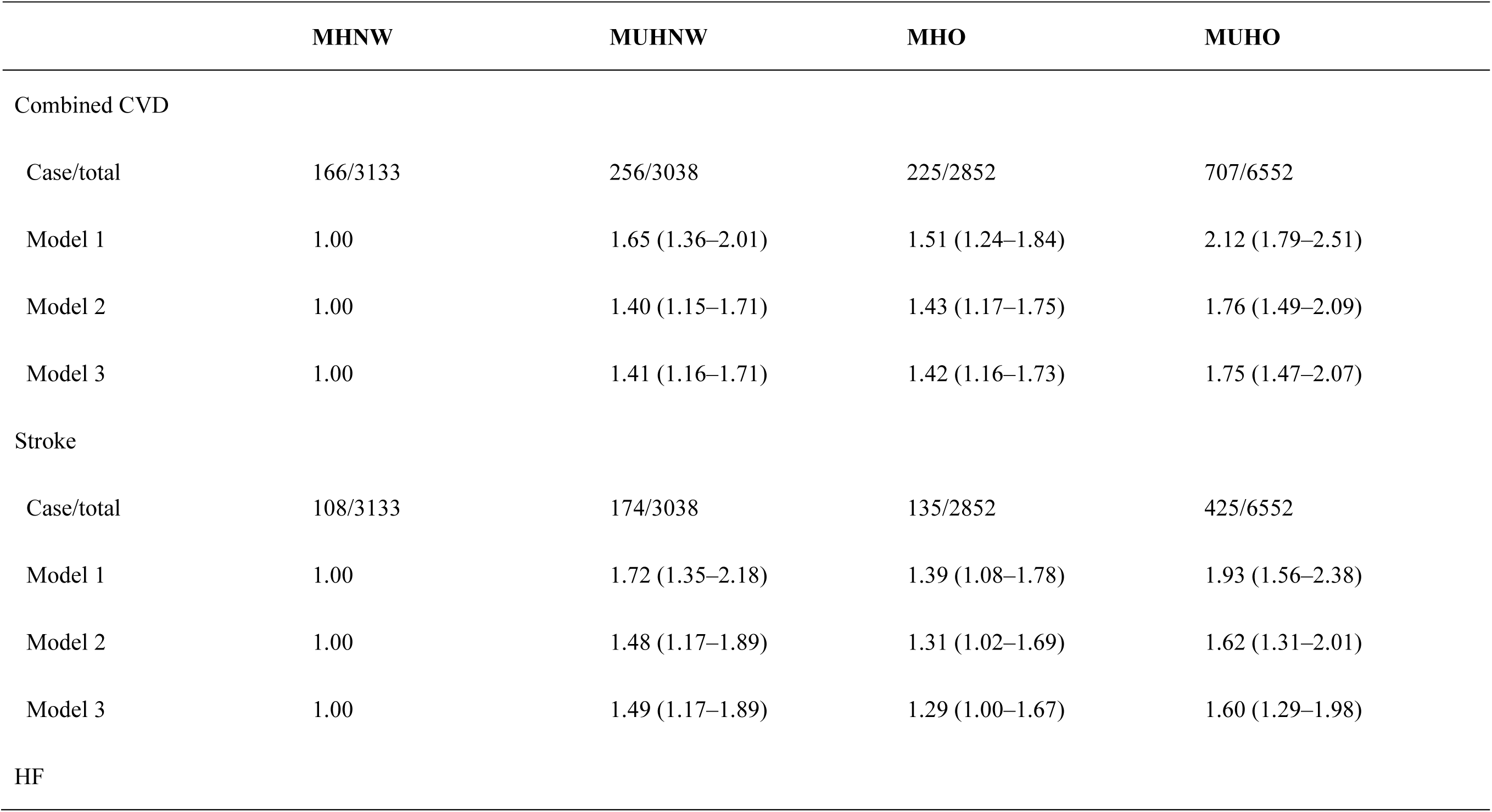
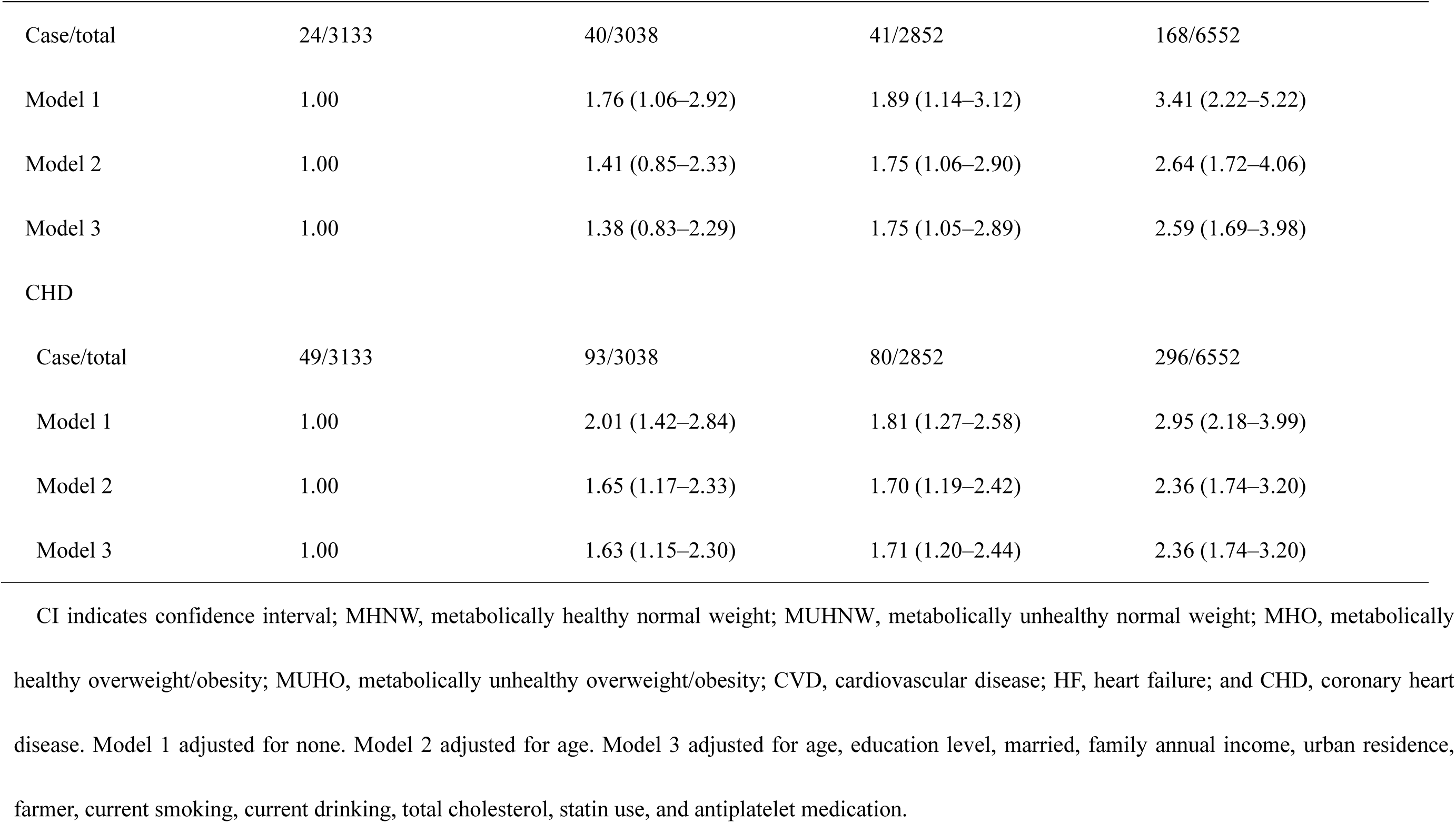
Hazard Ratios (95% CI) for Cardiovascular Outcomes Associated with Metabolic Weight Categories in Postmenopausal Women.

From sensitivity analyses (6 metabolic weight phenotypes), the fully adjusted HRs with 95% CI of the MUHNW [1.35 (1.14–1.60)], metabolically unhealthy overweight [1.49 (1.26–1.76)] and metabolically unhealthy obesity [1.71 (1.41–2.08)] were statistically significant for combined CVD compared with MHNW. The results for stroke and CHD were similar to those for combined CVD. In contrast to the results for combined CVD, stroke and CHD, metabolically healthy obesity and metabolically unhealthy overweight and obesity were significantly correlated with HF risk (**Table S1**).

### Additional Analyses

Among separate obesity (BMI and WC) and metabolic (SBP, DBP, TG, FBG, and HDL-C) measures, each stratified by their cut points as previous definitions, elevated WC (HR: 1.33; 95% CI: 1.16–1.51), elevated SBP (HR: 1.29; 95% CI: 1.13–1.47), and elevated DBP (HR: 1.22; 95% CI: 1.08–1.38) were significantly related to combined CVD after adjusting for covariates. In addition, elevated WC [1.34 (1.13–1.58)], elevated SBP [1.35 (1.13–1.60)] and elevated DBP [1.39 (1.19–1.62)] were associated with an increased risk of stroke. Moreover, elevated WC [1.53 (1.13–2.07)] and elevated TG [1.32 (1.02–1.70)] were risk factors for HF. In addition, elevated WC, TG and FBG were significantly associated with CHD, exhibiting HRs of 1.26 (1.01–1.56), 1.38 (1.15–1.66) and 1.26 (1.04–1.51), respectively (**Table 3**).

**Table 3.** Cox Regression of Cardiovascular Outcomes According to Separate Obesity and Metabolic Risk Factors Categorized by Cut Points.

| **Risk factor** | **Combined CVD** | **Stroke** | **HF** | **CHD** |
| --- | --- | --- | --- | --- |
| BMI ($\geq 24$ vs. 18.5–23.9 kg/m<sup>2</sup>) | 0.97 (0.85–1.11) | 0.83 (0.70–0.98) | 1.21 (0.89–1.63) | 1.22 (0.98–1.51) |
| WC ($\geq 85$ vs. $< 85$ cm) | 1.33 (1.16–1.51) | 1.34 (1.13–1.58) | 1.53 (1.13–2.07) | 1.26 (1.01–1.56) |
| SBP ($\geq 130$ vs. $< 130$ mm Hg) | 1.29 (1.13–1.47) | 1.35 (1.13–1.60) | 1.27 (0.94–1.72) | 1.15 (0.93–1.42) |
| DBP ($\geq 85$ vs. $< 85$ mm Hg) | 1.22 (1.08–1.38) | 1.39 (1.19–1.62) | 1.05 (0.80–1.38) | 1.10 (0.90–1.35) |
| TG ($\geq 1.7$ vs. $< 1.7$ mmol/L) | 1.10 (0.98–1.23) | 0.98 (0.85–1.14) | 1.32 (1.02–1.70) | 1.38 (1.15–1.66) |
| FBG ($\geq 5.6$ vs. $< 5.6$ mmol/L) | 1.12 (1.00–1.25) | 1.09 (0.95–1.26) | 1.11 (0.86–1.42) | 1.26 (1.04–1.51) |
| HDL-C ($< 1.3$ vs. $\geq 1.3$ mmol/L) | 1.03 (0.90–1.17) | 1.10 (0.93–1.29) | 1.03 (0.78–1.36) | 1.06 (0.86–1.29) |

BMI indicates body mass index; WC, waist circumference; SBP, systolic blood pressure; DBP, diastolic blood pressure; TG, triglyceride; FBG, fasting blood glucose; and HDL-C, high-density lipoprotein cholesterol. Adjusted for age, education level, marital status, family annual income, urban residence, farmer, current smoking, current drinking, total cholesterol, statin use, and antiplatelet medication.

According to **Table 4**, the risk of each endpoint was significantly positively related to the number of abnormal metabolic components. Compared with zero abnormal metabolic component, 3 + 4 abnormal metabolic components had the highest risk of combined CVD (HR: 1.96; 95% CI: 1.38–2.80), followed by 2 components (HR: 1.22; 95% CI: 1.05–1.41). Furthermore, with zero abnormal metabolic component as a reference, only 3 + 4, both 1 and 3 + 4, and the remaining sets showed increased risks of stroke, HF and CHD, respectively (all *P* for trend < 0.05).

**Table 4.** Cox Regression of Cardiovascular Outcomes According to the Number of Abnormal Metabolic Components.

| **Number of abnormal metabolic components** | **Combined CVD** | **Stroke** | **HF** | **CHD** |
| --- | --- | --- | --- | --- |
| 0 | 1.00 | 1.00 | 1.00 | 1.00 |
| 1 | 1.03 (0.91–1.16) | 0.99 (0.84–1.16) | 1.35 (1.02–1.78) | 1.24 (1.01–1.52) |
| 2 | 1.22 (1.05–1.41) | 1.20 (0.99–1.45) | 1.38 (0.99–1.93) | 1.58 (1.25–2.00) |
| 3 + 4 | 1.96 (1.38–2.80) | 1.93 (1.23–3.04) | 2.50 (1.23–5.06) | 2.16 (1.25–3.73) |
| *P* for trend | 0.0007 | 0.02 | 0.008 | <0.0001 |

CVD indicates cardiovascular disease; HF, heart failure; and CHD, coronary heart disease. Adjusted for age, education level, married, family annual income, urban residence, farmer, current smoking, current drinking, body mass index, waist circumference, total cholesterol, statin use, and antiplatelet medication.

From the results of subgroup analyses for age, education level, family annual income and residence (urban), we found no interactions among them (all *P* for interaction > 0.05) (**Table S2**).

## DISCUSSION

Our study suggested that in postmenopausal women from China, those with MUHNW, MHO and MUHO had a 41%, 42% and 75% greater risk of developing CVD than those with MHNW, respectively. Similar associations were observed for CHD. In addition, MUHNW participants had a higher risk of stroke than their MHO counterparts. The HF risk was significantly higher in subjects with MHO than MUHNW, indicating that obesity alone may contribute more to HF risk than metabolically unhealthy status in postmenopausal women. In addition, elevated WC was the most consistent individual metabolic/obesity predictor for cardiovascular outcomes. Risks of CVD were positively related to the number of abnormal metabolic components.

Consistent with previous cohort studies, the current research indicated that MHO and MUHNW individuals were at elevated risk of total CVD. In the China Health and Retirement Longitudinal Study of 7849 Chinese adults, Li et al. showed that MHO (HR: 1.33; 95% CI: 1.19–1.49) and MUHNW (HR: 1.29; 95% CI: 1.22–1.38) participants were at similarly greater risks of CVD during a mean follow-up of 3.67 years.^7^ Nevertheless, a meta-analysis comprising thirteen prospective studies in Western populations reported that MHO subjects were at 45%, but MUHNW subjects were at a much higher cardiovascular risk of 107% compared with MHNW subjects.^8^ However, this meta-analysis did not include Asian studies. Similarly, a Korean cohort including 514 866 participants followed for < 5 years suggested that MUHNW individuals were significantly more likely than MHO individuals to develop cardiovascular events,^9^ consistent with the results of a Taiwanese survey among 5358 subjects, but the associations of MHO were found to be stronger in women than in men in this survey.^10^ Different from our results, the Northeast China Rural Cardiovascular Health Study including 7472 participants aged ≥ 35 years demonstrated that MHO [1.48 (1.07–2.06)] instead of MUHNW [1.16 (0.82–1.65)] was significantly associated with combined CVD,^11^ in contrast to another study of the same cohort of 9345 residents indicating a significant association of MUHNW [1.63 (1.16–2.31)] with that rather than MHO [1.15 (0.67–1.99)] according to a novel criterion for the definition of metabolic health.^12^ However, none of the above studies focused on postmenopausal women, and our findings in this population suggest that both MHO and MUHNW were associated with an increased risk of combined CVD, with a slightly higher risk for MHO than for MUHNW. Furthermore, the present study demonstrated that the correlations of MUHNW with stroke were stronger than those of MHO, although the latter was still statistically significant, which was similar to the results of previous cohort investigations.^7, 22, 23^ Our study also suggested that the risk of HF was at a 75% increase in subjects with MHO, in line with the Health Improvement Network cohort of 3.5 million men and women reporting that those with MHO had a 96% higher risk than MHNW.^24^ However, this was inconsistent with another previous article of the Women’s Health Initiative including 19 412 postmenopausal women aged 50 to 79 years with findings that it was MUHNW but not MHO associated with an increased risk of HF.^13^

Interestingly, from the results of sensitivity analyses, we found that in contrast to metabolically unhealthy subjects across BMI categories, metabolically healthy subjects had no significant increase in the risk of combined CVD, which was consistent with the results of the Tehran Lipid and Glucose Study (TLGS) after a 12-year follow-up in 7167 participants.^14^ However, in other previous cohort studies,^25–27^ both metabolically healthy and metabolically unhealthy subjects categorized by weight were significantly associated with CVD compared with the reference MHNW, and interestingly, magnitudes of the risk among the 6 groups increased step by step. Similar results were observed for CVD and CHD in the Women’s Health Study of 25 626 women aged 45 years or older, although the associations were not significant in overweight women without metabolic syndrome.^28^ Such discrepancies can be partly explained by the relatively small sample sizes of metabolically healthy obese participants in our study (n = 490) and the TLGS (n = 147).

Previous studies showed that MHO individuals were more susceptible to subclinical carotid and coronary atherosclerosis.^29, 30^ This suggests that MHO is not a benign phenotype mainly due to obesity. A metabolically unhealthy status is easily transformed from insulin resistance and systematic inflammation induced by adipocyte tissue dysfunction and cytokine secretion and ultimately results in the development of CVD.^8^ Meanwhile, some normal-weight individuals were not necessarily metabolically healthy and had previously been reported to have metabolic abnormalities.^31^ Regardless of BMI, being metabolically unhealthy generally increases the risk of CVD events.^24^ Sex hormones, as effect regulators, can be involved in signaling through estrogen receptors, which play an important role in mediating cardiometabolic protective functions.^32^ Mechanistically, the adverse cardiometabolic effects of sex hormone deficiency that occur after menopause are due to impaired estrogen receptor signaling action, leading to significantly increased CVD risk.^33^ Studies have shown that increasing adiposity,^34^ insulin resistance, and inflammation^35^ are particularly caused by the loss of estrogen receptor α signaling, often prior to the development of atherosclerosis. Nevertheless, postmenopausal hormone replacement therapy is not recommended for primary or secondary CVD prevention.^36^ Of note, approximately 33%–52% of MHO women experience a conversion to metabolically unhealthy status in 6–20 years.^37^ Additionally, a BMI ≥ 40 kg/m^2^ and central adiposity characterized by a WC ≥ 88 cm were reported to be associated with a higher risk of mortality and sedentary behavior in postmenopausal women.^38^ High blood pressure is prevalent in women aged ≥ 50 years who are more likely to develop hypertension-related CVD than men.^39^ These findings warrant the need to improve lifestyles and diet for postmenopausal women as well as regularly assess related metabolic markers for those with WHO to delay the transition to MUHO, and it is necessary to enhance health surveillance in the MUNW population, as the status can still be correlated with an elevated risk of CVD.

Several limitations should be noted. First, our study only included postmenopausal women, and therefore, the results may not be applicable in younger people. Second, we did not measure visceral fat directly, which can better assess obesity. Third, although important confounders were adjusted in the multivariable model, we cannot guarantee excluding all possible confounding factors due to no measurement of variables such as physical activity and concentration of HbA1c. Last, metabolic weight groups were categorized only at baseline, as we did not repeat the measurement of obesity or metabolic components to see the effects of transitions among phenotypes and changes in risk factors over time on outcomes.

## CONCLUSIONS

Our study demonstrated that postmenopausal women with metabolically healthy overweight/obesity and metabolically unhealthy normal weight were at similar increased cardiovascular risk, which was much greater in those with metabolically unhealthy overweight/obesity. These findings indicate that metabolic abnormalities increase CVD risk in postmenopausal women with overweight/obesity and even normal weight.

## Acknowledgments

We thank all the participants included in this project.

## Supplementary Material

Data S1

## Nonstandard Abbreviations and Acronyms

MHNW: metabolically healthy normal weight
MUHNW: metabolically unhealthy normal weight
MHO: metabolically healthy overweight/obesity
MUHO: metabolically unhealthy overweight/obesity
China PEACE MPP: China Patient-centered Evaluative Assessment of Cardiac Events Million Persons Project

## REFERENCES

1. Roth GA, Mensah GA, Johnson CO, Addolorato G, Ammirati E, Baddour LM, Barengo NC, Beaton AZ, Benjamin EJ, Benziger CP, et al. Global Burden of Cardiovascular Diseases and Risk Factors, 1990-2019: Update From the GBD 2019 Study. J Am Coll Cardiol. 2020;76(25):2982–3021. doi:10.1016/j.jacc.2020.11.010

2. Vogel B, Acevedo M, Appelman Y, Bairey Merz CN, Chieffo A, Figtree GA, Guerrero M, Kunadian V, Lam CSP, Maas A, et al. The Lancet women and cardiovascular disease Commission: reducing the global burden by 2030. Lancet. 2021;397(10292):2385–2438. doi:10.1016/S0140-6736(21)00684-X

3. Virani SS, Alonso A, Aparicio HJ, Benjamin EJ, Bittencourt MS, Callaway CW, Carson AP, Chamberlain AM, Cheng S, Delling FN, et al. Heart Disease and Stroke Statistics-2021 Update: A Report From the American Heart Association. Circulation. 2021;143(8):e254-e743. doi:10.1161/CIR.0000000000000950

4. Lopez-Jimenez F, Almahmeed W, Bays H, Cuevas A, Di Angelantonio E, le Roux CW, Sattar N, Sun MC, Wittert G, Pinto FJ, et al. Obesity and cardiovascular disease: mechanistic insights and management strategies. A joint position paper by the World Heart Federation and World Obesity Federation. Eur J Prev Cardiol. 2022;29(17):2218–2237. doi:10.1093/eurjpc/zwac187

5. Kivimaki M, Kuosma E, Ferrie JE, Luukkonen R, Nyberg ST, Alfredsson L, Batty GD, Brunner EJ, Fransson E, Goldberg M, et al. Overweight, obesity, and risk of cardiometabolic multimorbidity: pooled analysis of individual-level data for 120 813 adults from 16 cohort studies from the USA and Europe. Lancet Public Health. 2017;2(6):e277–e285. doi:10.1016/S2468-2667(17)30074-9

6. Silveira Rossi JL, Barbalho SM, Reverete de Araujo R, Bechara MD, Sloan KP, Sloan LA. Metabolic syndrome and cardiovascular diseases: Going beyond traditional risk factors. Diabetes Metab Res Rev. 2022;38(3):e3502. doi:10.1002/dmrr.3502

7. Li H, He D, Zheng D, Amsalu E, Wang A, Tao L, Guo J, Li X, Wang W, Guo X. Metabolically healthy obese phenotype and risk of cardiovascular disease: Results from the China Health and Retirement Longitudinal Study. Arch Gerontol Geriatr. 2019;82:1–7. doi:10.1016/j.archger.2019.01.004

8. Eckel N, Meidtner K, Kalle-Uhlmann T, Stefan N, Schulze MB. Metabolically healthy obesity and cardiovascular events: A systematic review and meta-analysis. Eur J Prev Cardiol. 2016;23(9):956–966. doi:10.1177/2047487315623884

9. Cho YK, Kang YM, Yoo JH, Lee J, Park JY, Lee WJ, Kim YJ, Jung CH. Implications of the dynamic nature of metabolic health status and obesity on risk of incident cardiovascular events and mortality: a nationwide population-based cohort study. Metabolism. 2019;97:50–56. doi:10.1016/j.metabol.2019.05.002

10. Yeh TL, Hsu HY, Tsai MC, Hsu LY, Hwang LC, Chien KL. Association between metabolically healthy obesity/overweight and cardiovascular disease risk: A representative cohort study in Taiwan. PLoS One. 2021;16(2):e0246378. doi:10.1371/journal.pone.0246378

11. Guo X, Li Z, Zhou Y, Yu S, Yang H, Sun G, Zheng L, Afzal J, Liu Y, Sun Y. The effects of transitions in metabolic health and obesity status on incident cardiovascular disease: Insights from a general Chinese population. Eur J Prev Cardiol. 2021;28(11):1250–1258. doi:10.1177/2047487320935550

12. Li Q, Wang P, Ma R, Guo X, Sun Y, Zhang X. A novel criterion of metabolically healthy obesity could effectively identify individuals with low cardiovascular risk among Chinese cohort. Front Endocrinol (Lausanne). 2023;14:1140472. doi:10.3389/fendo.2023.1140472

13. Cordola Hsu AR, Xie B, Peterson DV, LaMonte MJ, Garcia L, Eaton CB, Going SB, Phillips LS, Manson JE, Anton-Culver H, et al. Metabolically Healthy/Unhealthy Overweight/Obesity Associations With Incident Heart Failure in Postmenopausal Women: The Women’s Health Initiative. Circ Heart Fail. 2021;14(4):e007297. doi:10.1161/CIRCHEARTFAILURE.120.007297

14. Mirzaei B, Abdi H, Serahati S, Barzin M, Niroomand M, Azizi F, Hosseinpanah F. Cardiovascular risk in different obesity phenotypes over a decade follow-up: Tehran Lipid and Glucose Study. Atherosclerosis. 2017;258:65–71. doi:10.1016/j.atherosclerosis.2017.02.002

15. Lu J, Xuan S, Downing NS, Wu C, Li L, Krumholz HM, Jiang L. Protocol for the China PEACE (Patient-centered Evaluative Assessment of Cardiac Events) Million Persons Project pilot. BMJ Open. 2016;6(1):e010200. doi:10.1136/bmjopen-2015-010200

16. Li X, Wu C, Lu J, Chen B, Li Y, Yang Y, Hu S, Li J. Cardiovascular risk factors in China: a nationwide population-based cohort study. Lancet Public Health. 2020;5(12):e672–e681. doi:10.1016/S2468-2667(20)30191-2

17. Lu J, Lu Y, Yang H, Bilige W, Li Y, Schulz W, Masoudi FA, Krumholz HM. Characteristics of High Cardiovascular Risk in 1.7 Million Chinese Adults. Ann Intern Med. 2019;170(5):298–308. doi:10.7326/M18-1932

18. Chinese Nutrition Society Obesity P, Control S, Chinese Nutrition Society Clinical Nutrition S, Chinese Preventive Medicine Association Behavioral Health S, Chinese Preventive Medicine Association S, Health S. [Expert Consensus on Obesity Prevention and Treatment in China]. Zhonghua Liuxingbingxue Zazhi. 2022;43(5):609–626. doi:10.3760/cma.j.cn112338-20220402-00253

19. Bao Y, Lu J, Wang C, Yang M, Li H, Zhang X, Zhu J, Lu H, Jia W, Xiang K. Optimal waist circumference cutoffs for abdominal obesity in Chinese. Atherosclerosis. 2008;201(2):378–384. doi:10.1016/j.atherosclerosis.2008.03.001

20. Alberti KG, Eckel RH, Grundy SM, Zimmet PZ, Cleeman JI, Donato KA, Fruchart JC, James WP, Loria CM, Smith SC, Jr. Harmonizing the metabolic syndrome: a joint interim statement of the International Diabetes Federation Task Force on Epidemiology and Prevention; National Heart, Lung, and Blood Institute; American Heart Association; World Heart Federation; International Atherosclerosis Society; and International Association for the Study of Obesity. Circulation. 2009;120(16):1640–1645. doi:10.1161/circulationaha.109.192644

21. Sattar N, Rawshani A, Franzén S, Rawshani A, Svensson AM, Rosengren A, McGuire DK, Eliasson B, Gudbjörnsdottir S. Age at Diagnosis of Type 2 Diabetes Mellitus and Associations With Cardiovascular and Mortality Risks. Circulation. 2019;139(19):2228–2237. doi:10.1161/circulationaha.118.037885

22. Zhou Y, Zhang X, Zhang L, Li Z, Wu Q, Jin Z, Chen S, He D, Wu S, Zhu Y. Increased Stroke Risk in Metabolically Abnormal Normal Weight: a 10-Year Follow-up of 102,037 Participants in China. Transl Stroke Res. 2021;12(5):725–734. doi:10.1007/s12975-020-00866-1

23. Lee HJ, Choi EK, Lee SH, Kim YJ, Han KD, Oh S. Risk of ischemic stroke in metabolically healthy obesity: A nationwide population-based study. PLoS One. 2018;13(3):e0195210. doi:10.1371/journal.pone.0195210

24. Caleyachetty R, Thomas GN, Toulis KA, Mohammed N, Gokhale KM, Balachandran K, Nirantharakumar K. Metabolically Healthy Obese and Incident Cardiovascular Disease Events Among 3.5 Million Men and Women. J Am Coll Cardiol. 2017;70(12):1429–1437. doi:10.1016/j.jacc.2017.07.763

25. Fan J, Song Y, Chen Y, Hui R, Zhang W. Combined effect of obesity and cardio-metabolic abnormality on the risk of cardiovascular disease: a meta-analysis of prospective cohort studies. Int J Cardiol. 2013;168(5):4761–4768. doi:10.1016/j.ijcard.2013.07.230

26. Gao M, Lv J, Yu C, Guo Y, Bian Z, Yang R, Du H, Yang L, Chen Y, Li Z, et al. Metabolically healthy obesity, transition to unhealthy metabolic status, and vascular disease in Chinese adults: A cohort study. PLoS Med. 2020;17(10):e1003351. doi:10.1371/journal.pmed.1003351

27. Wang W, He J, Hu Y, Song Y, Zhang X, Guo H, Wang X, Keerman M, Ma J, Yan Y, et al. Comparison of the Incidence of Cardiovascular Diseases in Weight Groups with Healthy and Unhealthy Metabolism. Diabetes Metab Syndr Obes. 2021;14:4155–4163. doi:10.2147/DMSO.S330212

28. Song Y, Manson JE, Meigs JB, Ridker PM, Buring JE, Liu S. Comparison of usefulness of body mass index versus metabolic risk factors in predicting 10-year risk of cardiovascular events in women. Am J Cardiol. 2007;100(11):1654–1658. doi:10.1016/j.amjcard.2007.06.073

29. Itoh H, Kaneko H, Kiriyama H, Yoshida Y, Nakanishi K, Mizuno Y, Daimon M, Morita H, Yatomi Y, Yamamichi N, et al. Effect of Metabolically Healthy Obesity on the Development of Carotid Plaque in the General Population: A Community-Based Cohort Study. J Atheroscler Thromb. 2020;27(2):155–163. doi:10.5551/jat.48728

30. Chang Y, Kim BK, Yun KE, Cho J, Zhang Y, Rampal S, Zhao D, Jung HS, Choi Y, Ahn J, et al. Metabolically-healthy obesity and coronary artery calcification. J Am Coll Cardiol. 2014;63(24):2679–2686. doi:10.1016/j.jacc.2014.03.042

31. St-Onge MP, Janssen I, Heymsfield SB. Metabolic syndrome in normal-weight Americans: new definition of the metabolically obese, normal-weight individual. Diabetes Care. 2004;27(9):2222–2228. doi:10.2337/diacare.27.9.2222

32. Clegg D, Hevener AL, Moreau KL, Morselli E, Criollo A, Van Pelt RE, Vieira-Potter VJ. Sex Hormones and Cardiometabolic Health: Role of Estrogen and Estrogen Receptors. Endocrinology. 2017;158(5):1095–1105. doi:10.1210/en.2016-1677

33. Ouyang P, Michos ED, Karas RH. Hormone replacement therapy and the cardiovascular system lessons learned and unanswered questions. J Am Coll Cardiol. 2006;47(9):1741–1753. doi:10.1016/j.jacc.2005.10.076

34. Rettberg JR, Yao J, Brinton RD. Estrogen: a master regulator of bioenergetic systems in the brain and body. Front Neuroendocrinol. 2014;35(1):8–30. doi:10.1016/j.yfrne.2013.08.001

35. Das SK, Ma L, Sharma NK. Adipose tissue gene expression and metabolic health of obese adults. Int J Obes (Lond). 2015;39(5):869–873. doi:10.1038/ijo.2014.210

36. Shufelt CL, Manson JE. Menopausal Hormone Therapy and Cardiovascular Disease: The Role of Formulation, Dose, and Route of Delivery. J Clin Endocrinol Metab. 2021;106(5):1245–1254. doi:10.1210/clinem/dgab042

37. Eckel N, Li Y, Kuxhaus O, Stefan N, Hu FB, Schulze MB. Transition from metabolic healthy to unhealthy phenotypes and association with cardiovascular disease risk across BMI categories in 90 257 women (the Nurses’ Health Study): 30 year follow-up from a prospective cohort study. Lancet Diabetes Endocrinol. 2018;6(9):714–724. doi:10.1016/S2213-8587(18)30137-2

38. El Khoudary SR, Aggarwal B, Beckie TM, Hodis HN, Johnson AE, Langer RD, Limacher MC, Manson JE, Stefanick ML, Allison MA, et al. Menopause Transition and Cardiovascular Disease Risk: Implications for Timing of Early Prevention: A Scientific Statement From the American Heart Association. Circulation. 2020;142(25):e506–e532. doi:10.1161/CIR.0000000000000912

39. Nappi RE, Simoncini T. Menopause transition: a golden age to prevent cardiovascular disease. Lancet Diabetes Endocrinol. 2021;9(3):135–137. doi:10.1016/S2213-8587(21)00018-8

